# Health care seeking behaviour for common menstrual problems and their determinants in Adolescent girls at rural setting in Central India: A mix-method study

**DOI:** 10.1101/2023.08.01.23293531

**Authors:** S Patel, B Patel, K Pushpalatha, A Halder, JN Modi, B Singh

**Affiliations:** Department of Obstetrics and Gynecology, AIIMS Bhopal; Department of Pediatric Trauma and Emergency Medicine, AIIMS Bhopal

**Keywords:** Menstruation, Menstrual health problems, menstrual hygiene, Adolescent girls, health seeking behaviour

## Abstract

**Introduction:** Menstrual problems among adolescent females are common and a significant source of morbidity in this population.This study was to collect data on the different health seeking behaviours used by adolescents and their determinants for common menstrual illness in rural population in central India. Collected information may be vital for the development of appropriate adolescent healthcare system.

**Aims & Objectives:** To assess the health care seeking behaviour and factors determinants in adolescent girl/ their parents for menstrual problems in rural areas ofBhopal.

**Material & Methods:** This cross sectional study was done in nearby (within 30km radius of AIIMS Bhopal) schools villages on adolescent girls (12-19 years) after excluding the chronic illnesses. By using multistage cluster sampling 10 village schools were selected randomly. After taking informed consent from the head of the school 40 girls from each school were enrolled. The information were collected using a pre tested, semi structured questionnaire from all 400 participants. Statistical analysis was done by using IBM-SPSS version 22 software.

**Results:** The mean age of the girls in the study was 17±1.4 years. In our study 27% of adolescent girls suffered from menstrual problems with dysmenorrhoea (48%) was the most frequent complaint out of which nearly one-third of adolescent girls sought treatment. Majority (67%) girls didn’t know about menstruation prior to menarche. Social barriers/ restrictions, shame and misconceptions with the menstruations were commonly reported in these girls.

**Conclusion:** Adolescent girls are reluctant to seek medical advice and with little knowledge on adolescent’s health-seeking behavior towards the menstrual problems leading to delay in the diagnosis and treatment. Adolescent health educational programmes in the society and in the schools help the girls toimprove the treatment-seeking for menstrual problems, specifically in rural areas.

## Introduction

The World Health Organization (WHO) defines adolescents as people who are currently between the ages of 10 and 19 years old {1}. Secondary sexual traits, sexual development, and reproductive capacity are often achieved at this age. Ovulation and the beginning of menstruation both begin during this time period in girls {2}. This time period is also frequently accompanied with menstrual problems and other related morbidities {3}.

Different cultures have always held a variety of views around menstruation, and this has never changed. Today, there is a greater acceptance of menstruation than there was in the past; nonetheless, disparities in mentality still exist amongst different civilizations {4}. There are disparities between countries, civilizations, religions, and ethnic groups regarding practises related to menstruation. In many low-income nations, women and girls are restricted in mobility and conduct during menstruation due to their “impurity” during menstruation. Menstruation is still associated with a number of cultural taboos, as well as emotions of shame and uncleanliness in many parts of the world. Menstruation is still a mother and daughter secret in many families today. It is not openly discussed {5}.Menstruation is regarded a natural event in India, a gift from God, and is considered vital. Women’s perceptions of menstruation differ among cultures and religions. There are various taboos, associated with menstruation in India. Young women in India have little understanding and numerous misconceptions regarding menstruation before and after menarche. This frequently results in excessive dread, worry, and undesired behaviours. Menstruation knowledge and habits are influenced by socioeconomic factors as well {6}.

Due to later childbearing and fewer children, the number of women in developing nations, including India, who have regular menstruation periods is increasing. However, many people lack the economic and social resources to manage menstrual sanitation adequately. Young women from low-income homes are particularly vulnerable in this regard. Furthermore, understanding young women’s menstrual knowledge and practice’s is critical for developing suitable education programmes {6}.

In low and middle-income nations, like India, there is very little social and health research on menstruation disorders. There is also a scarcity of study on menstruation as a social and cultural phenomenon, as well as the technical and sanitary aspects of sanitary protection in varied socioeconomic circumstances. The reason for this could be that menarche and menstruation are taboo topics that are rarely discussed, even between mother and daughter {7}.

Taking the above scenario, this study was designed to understand health care seeking behaviour and knowledge regarding menstruation in young & adolescent girls residing in rural areas of Central India.

## Methodology

This is a cross-sectional study which included 400 female adolescent participants who have reached menarche. By using multistage cluster sampling 10 village schools were selected randomly. 40 girls were enrolled from each village. Informed accent was taken from participants and informed written consent was obtained from head of the school and parents. Ethical clearance was taken from Institutional human ethics committee of our institute.

### Inclusion criteria

- Adolescent girls in the age group of 10-19 years in the selected villages
- Must have reached menarche.
- Ability to understand and respond to questionnaires

### Exclusion criteria

- Pregnant and lactating adolescent mothers
- Adolescent females who haven’t reached menarche.

The information was collected using a pre tested, semi structured questionnaire from all 400 participants. Statistical analysis was done by using IBM-SPSS version 22 software.

## Results

Mean age of participants in our study, 15±1.3years. Menstrual abnormalities were reported by 27% of the participants. Of those, dysmenorrhea was the most common complaint, affecting 48% of adolescent girls. However, only 33% of adolescent girls sought treatment for their symptoms. Prior to reaching menarche, the majority of girls (67%) were unaware that they had periods. It was typical for these girls to report experiencing social barriers or restrictions, feelings of shame, and misconceptions around menstruation.

**Table 1.** illustrates Sociodemographic characteristics of participants included in the study.

| SN | Variables |  | No (%) |
| --- | --- | --- | --- |
| 1 | Age | Early Adolescent<br>10-14 years | 168 (42%) |
|  |  | Late Adolescents<br>15-19 years | 232 (58%) |
| | | Mean Age | $15 \pm 1.3$ |
| 2 | Religion | Hindu | 301 (75.25%) |
|  |  | Muslim | 52 (13.0%) |
|  |  | Christian | 21 (05.25%) |
|  |  | Other | 26 (06.50%) |
| 3 | Socio economic status | Low | 196 (49.0%) |
|  |  | Middle | 153 (38.25%) |
|  |  | High | 51 (12.75%) |
| 4 | Type of Family | Nuclear | 163 (40.75%) |
|  |  | Joint | 237 (59.25%) |
| 5 | Health Insurance | Yes | 36 (09.0%) |
|  |  | No | 364 (91.0%) |
| 6 | Toilet Availability | In Home | 306 (76.5%) |
|  |  | Nearby | 66 (16.5%) |
|  |  | Not Available | 28 (07.0%) |

**Table 2.**
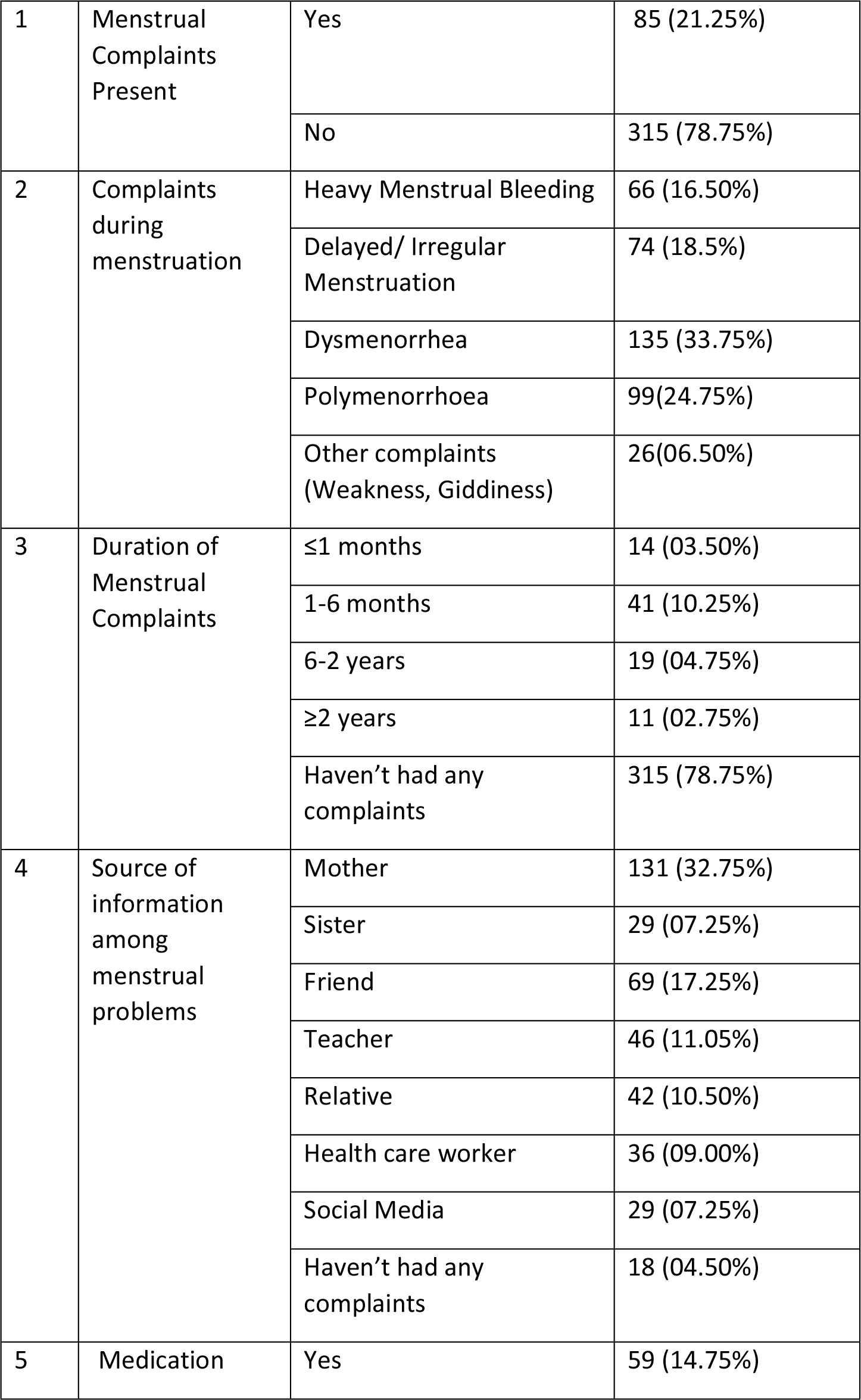
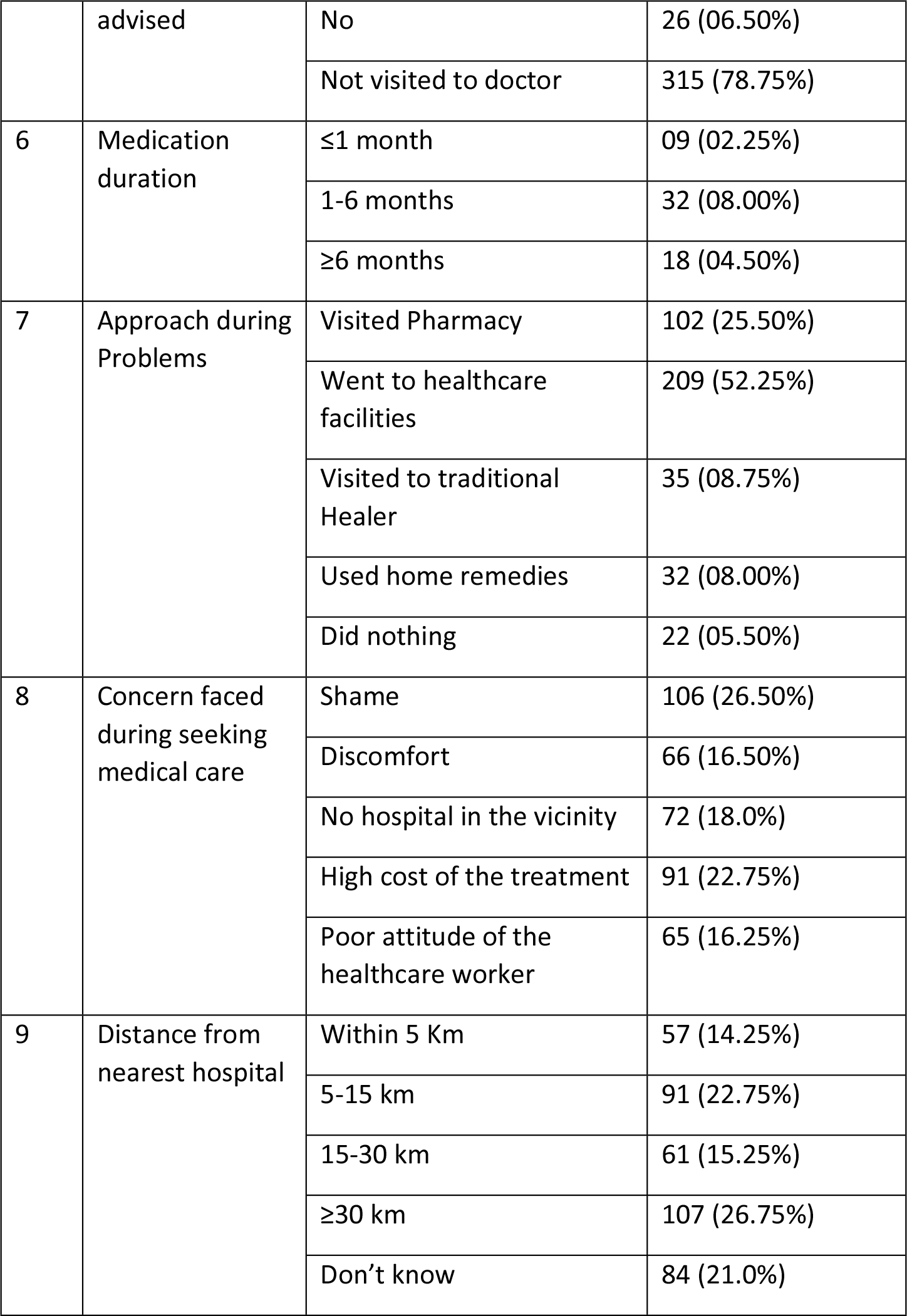
illustrates Common menstrual complaints and healthcare seeking behaviour among participants.

**Table 3.**
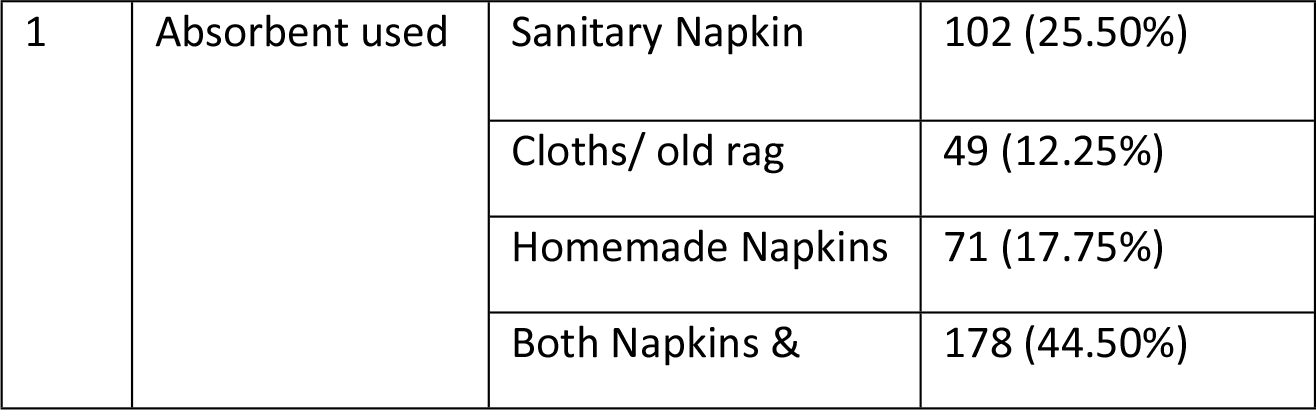
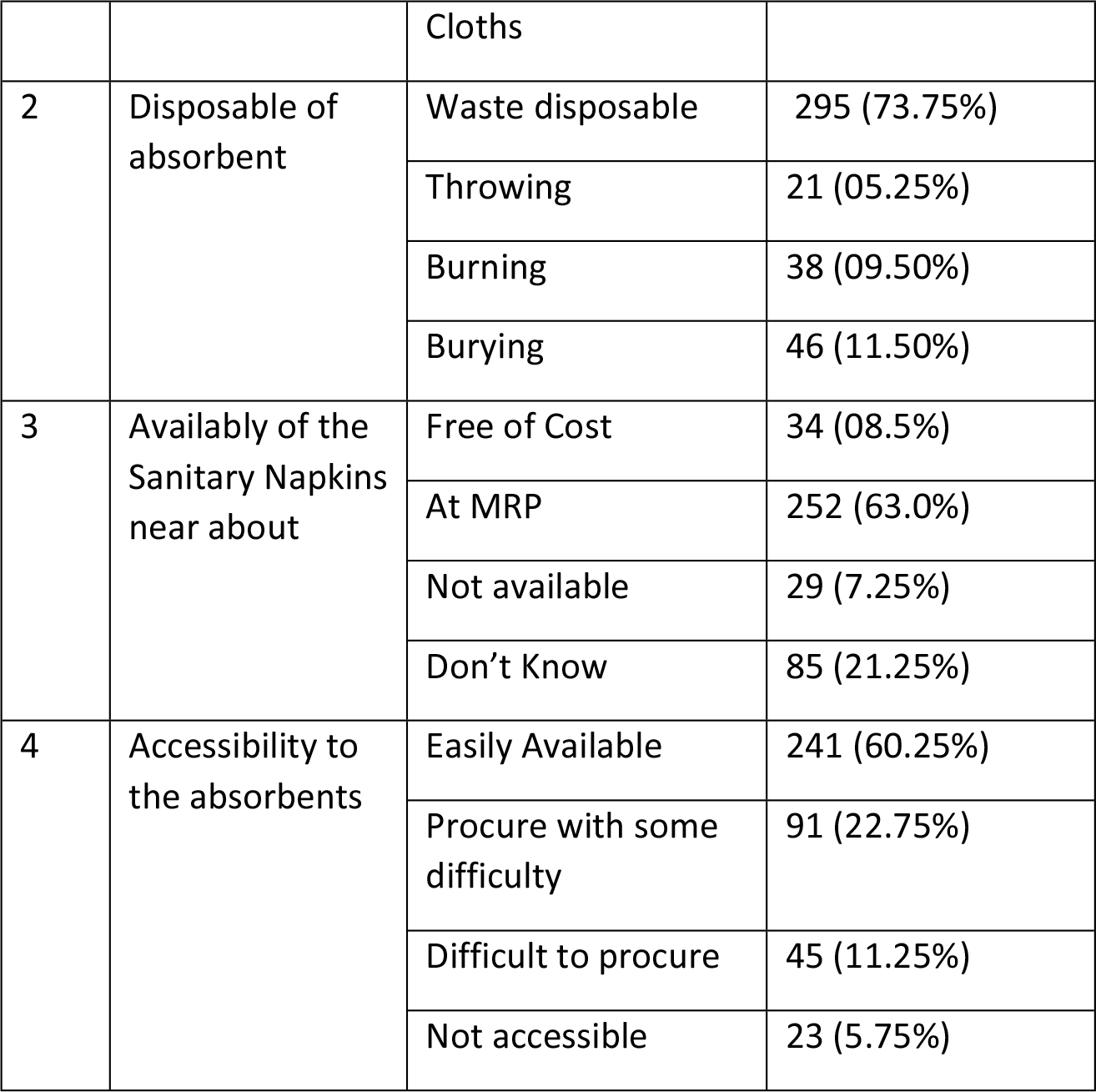
Menstrual hygiene practice and their determinants in the study.

## Discussion

The onset of menstruation is an unavoidable aspect of a girl’s life and, more importantly, an essential marker of healthy physical, physiological, and functional functioning.

The purpose of this study is to shed light on the menstrual condition of adolescent girls who live in rural areas close to Bhopal as well as their health seeking behaviour in terms of menstrual health. In our study, only 21.25% participants reported menstrual abnormalities; among these abnormalities, oligomenorrhea was most prevalent (8%) among participants followed by dysmenorrhea (6.5%), heavy menstrual bleeding (5.25%) and other complaints (1.5%) such as weakness & giddiness etc.

Most participants (32.75%) in our study reported that they discussed menstruation related to issues with their mothers followed by friend (17.25%), teacher (11.5%)& sisters (7.25%)which is in accordance to the findings by Nair et al (2007)& Tiwari et al (2006) {8, 9}.

Despite reporting menstrual abnormalities 78.75% of the participants did not seek any medical treatment. Participants reported multiple reasons for not seeking medical care. Most participants (26.5%) felt shameful and discomfort was reported by 16.5%. Lack of proper medical facilities and poor attitude of healthcare worker also effected health care seeking attitude among 18% and 16.25% of participants. Cost of treatment was also reported to be a barrier in seeking healthcare advises among 22.75% of the participants.Kabir et al (2014) also reported seeking healthcare can be difficult and time-consuming process for teens to seek treatment for gynaecological disorders {10}. It primarily depends on the individual’s level of ease and familiarity with the service providers, inaddition to the individual’s access to the health services.Previous studies have also reported that in rural areas, stigma related to gynaecological morbidities may be one reason for the lower treatment of gynaecological morbidities among adolescents. Moreover, in rural areas, health care services may be too far from home {11}.

Most participants in our study (44.5%) stated to use both sanitary napkin & cloth during their menstrual cycle; only 25.5% reported to use only sanitary napkin. Similar findings were reported by Thakur et al (2014) where 74% of participants in their study reported to use both napkin and cloth {12}.

Study by Lohani et al (2014) reported the increase in usage of sanitary napkins from 14.7% to 27% can be due to availability of sanitary napkin with ASHA at local level {13}. Ease of availability and cost of sanitary napkins majorly influence the choice of menstruating women in rural India. In a reviewEjik at el (2016) I t appeared that financial concerns were the primary motivation for using cloths rather than pads; other motivations were difficulties associated with disposal, ignorance of pads, or personal choice {14}.

Studies have also reported the usage of reusable cloth during menstruation by females belonging to low socio-economic backgrounds. According to the result research by Thakur et al (2014), just 25% of young women reused their garments (either alone or along with sanitary napkins). It is obvious that these families are struggling socioeconomically, and as a result, they are unable to afford the expensive sanitary napkins that they require. But it is absolutely necessary that the used cloths be washed in the appropriate manner.

In a study conducted in Rajasthan {15} and Delhi {8}, the researchers found that the vast majority of the young girls reused old fabric and homemade napkins, while only a very small percentage of them utilised cotton wool or sanitary napkins. The most common and economical form of protection worn during menstruation is made of cloth. The majority of women living in slums and rural areas make use of a wide variety of clothing, including that which has been worn out, torn, or otherwise disregarded by society. The difficulty to purchase ready-made sanitary napkins due to their high cost and the lack of availability in rural regions were the primary factors that led to the use of homemade sanitary napkins. Lack of information can lead to various unsanitary activities, including but not limited to: reusing the same cloth again and again without adequate cleaning; ignoring health problems; attempting to manage the challenges faced during menstruation on their own; and so on. During their periods, these preteen and teenage females are usually confronted with a variety of challenges and limitations{12}.

## Limitations

In retrospect, the researchers recognise that there were caveats to their study. Due to coverage limitations, we can only speak to people from middle-to-lower socioeconomic backgrounds in urban areas. Minorities and people living in rural areas were left out. Majority of the people there identified as Hindu. It was also impossible to study the perspectives of boys and adult men. In order to have a deeper understanding of these difficulties, it would be beneficial to conduct interviews both genders.

## Conclusion

The importance, urgency, and underappreciated requirement of disseminating accurate information to the entire community, particularly adolescent and young females, is reaffirmed in this research. With the right information, they can ditch harmful cultural norms and restrictions about menstruation and instead use safe, hygienic methods. This requires the formulation and implementation of appropriate policies, which can be a component of broader health and community development agendas. Family life/sexual education in schools should include discussions about the physiology of the menstrual cycle, the role it plays in fertility, and the fact that menstruation is a clean and normal part of a woman’s life. The healthcare industry, and the public health system in particular, needs to take the initiative.

## Data Availability

Whole data of study is available in the manuscript. There is no need to add the data elsewhere.

## References

1. World Health Organization. WHO factsheets on women’s health. http://www.who.int/mediacentre/factsheets/fs334/en/ (2009). Accessed 1 Jan 2021.

2. Talbert LM, Hammond MG, Grof T, Udry JR. Relationship of age and pubertal development to ovulation in adolescent girls. Obstet Gynecol. 1985;66:542–4.

3. Carlson LJ, Shaw ND. Development of ovulatory menstrual cycles in adolescent girls. J PediatrAdolGynec. 2019;32:249–53.

4. Cronje HS, Kritzinger IE. Menstruation: symptoms, management and attitudes in university students. Int J GynaecolObstet (1991) 35(2):147–50. doi:10.1016/0020-7292(91)90818-P

5. Bhatt R, Bhatt M. Perceptions of Indian women regarding menstruation. Int J GynaecolObstet (2005) 88(2):164–7. doi:10.1016/j.ijgo.2004.10.008

6. Mahon T, Fernandes M. Menstrual hygiene in South Asia: a neglected issue for WASH (water, sanitation and hygiene) programmes. GendDev (2010) 18:99–113. doi:10.1080/13552071003600083

7. Drakshayani Devi K, Venkata Ramaiah P. A study on menstrual hygiene among rural adolescent girls. Indian J Med Sci (1994) 48(6):139–43. Singh AJ. Place of menstruation in the reproductive lives of women of rural North India. Indian J Community Med (2006) 31(3):10–4. doi:10.4103/0970-0218.54923

8. Nair P, Grover V,Kannan A. Awareness and practices of menstruation and pubertal changes amongst unmarried female adolescents in a rural area of East Delhi. Indian J Community Med (2007) 32:156–7. doi:10.4103/0970-0218.35668

9. Tiwari H, Oza UN, Tiwari R. Knowledge, attitudes and beliefs about menarche of adolescent girls in Anand district, Gujarat. East Mediterr Health J (2006) 12(3–4):428–33

10. Kabir, H., Saha, N. C., Wirtz, A. L., & Gazi, R. (2014). Treatment-seeking for selected reproductive health problems: behaviours of unmarried female adolescents in two low-performing areas of Bangladesh. Reproductive Health, 11(1), 1–7.

11. Kinkor, M. A., Padhi, B. K., Panigrahi, P., & Baker, K. K. (2019). Frequency and determinants of health care utilization for symptomatic reproductive tract infections in rural Indian women: A cross-sectional study. Plos one, 14(12), e0225687.

12. Thakur, H., Aronsson, A., Bansode, S., StalsbyLundborg, C., Dalvie, S., & Faxelid, E. (2014). Knowledge, practices, and restrictions related to menstruation among young women from low socioeconomic community in Mumbai, India. Frontiers in public health, 2, 72.

13. Lohani, P. (2019). Prevalence and determinants of menstrual disorders and napkin usage among women in India using DLHS-4 data. Journal of family medicine and primary care, 8(6), 2106.

14. Van Eijk, A. M., Sivakami, M., Thakkar, M. B., Bauman, A., Laserson, K. F., Coates, S., & Phillips-Howard, P. A. (2016). Menstrual hygiene management among adolescent girls in India: a systematic review and meta-analysis. BMJ open, 6(3), e010290.

15. Khanna A, GoyalR S,Bhawsar R. Menstrual practices and reproductive problems: a study of adolescent girls in Rajasthan. J Health Manage (2005) 7(1):91–107. doi:10.1177/097206340400700103

